# Investigating the relationship between serum ACE 2 level and COVID-19 patients’ prognosis: a cross-sectional study

**DOI:** 10.1101/2021.05.02.21256329

**Authors:** Parsa Mohammadi, Hesam Aldin Varpaei, Arash Seifi, Sepideh Zahak Miandoab, Saba Beiranvand, Sahar Mobaraki, Mostafa Mohammadi

## Abstract

**Background:** The only known receptor for this virus in the human body is ACE2, the same known receptor for the SARS virus.

**Material and Method:** In this single-center cross-sectional study, 38 hospitalized adult (≥18 years) patients with laboratory-confirmed COVID-19 were identified in the infectious disease ward in Imam Khomeini hospital complex. The study also has been approved in ethics committee of Tehran University of medical sciences with ethic code: 99/11/101/16529. Data were analyzed using SPSS 25. p < 0.05 was considered statistically significant when a two-tailed test was performed.

**Result:** Among the 38 patients, the mean age was 64.13 years, 52.6% were male, 42% were PCR test positive and 39.5% was expired. The most common presenting symptoms were cough (80%), fever (75.5%), dyspnea (60.5%), myalgias (35.8%), diarrhea (20%), and nausea and vomiting (15%). There were not any significant differences between expired and discharged group in terms of serum ACE2 level. Results were similar between discharged and expired patients in the subgroup analysis of 38 patients.

**Conclusion:** It seems that serum ACE 2 level is not correlated with COVID-19 patients’ prognosis. However, it seems that more researches are required to confirm supposed association between serum ACE2 level and inflammatory biomarkers, clinical outcome, and patient’s survival.

## Introduction

The SARS-CoV-2 virus, the causative agent of the COVID-19 pandemic [1], is now a global problem and has infected more than 180 countries, including Iran. The only known receptor for this virus in the human body is ACE2, the same known receptor for the SARS virus [2]. The spike protein binds to the ACE2 receptor on the surface of human cells, then enters the cell and multiplies [3]. Because the ACE2 gene is most abundantly expressed in the heart, lung, and kidney tissues [4], many of the symptoms and problems of patients with COVID-19 are related to the virus interacting with the ACE2 protein, including cardiovascular and pulmonary problems [5, 6]. There are many suggestions for treating the new coronavirus, lots of these pieces of evidence are related to the interaction of ACE2 with the viral Spike protein; Such as hydroxychloroquine, which alters a portion of the ACE2 molecule and prevents it from binding to the virus [7], blocking the ACE2 receptor, or using a solution of ACE2 to bind to and neutralize the viral spike protein [6]. Also, changes in ACE2 in heart disease [8], kidney [9], diabetes [10] and in older ages [11] could be the reason for the greater vulnerability of this group of patients against SARS-CoV-2 virus.

Because SARS-CoV-2 enters host cells by binding to the angiotensin-converting enzyme (ACE2), the role of the renin-angiotensin-aldosterone (RAAS) system and immunosuppressants, or RAAS, in COVID-19 is controversial yet [12,13]. It seems that ACE2 is like a double-edged sword in COVID-19 infection. On the one hand, age, comorbidities, or treatment with renin-angiotensin-aldosterone system inhibitors may alter ACE2 expression and increase patients’ sensitivity to SARS-CoV-2 [14,15]. On the other hand, inhibition of the renin-angiotensin-aldosterone system or increased plasma ACE2 expression may degrade angiotensin II and reduce its destructive effects on ARDS in COVID-19 [16]. Thus, SARS-CoV-2 infection may alter ACE2 expression, thereby modulating the levels of angiotensin II and aldosterone [17].

All of this information indicates the importance of ACE2 levels monitoring in COVID-19 patients. Measurements in this study are performed using the sACE2 ELISA kit. By comparing serum ACE2 levels in newly admitted patients with sACE2 levels within 48 hours of the same patients, the value of sACE2 levels in determining the prognosis of COVID-19 patients can be determined. Therefore, the aim of this study was to use serum ACE2 levels to determine the prognosis of COVID-19 patients

## Methods and Materials

In this single-center cross-sectional study, 38 hospitalized adult (≥18 years) patients with laboratory-confirmed COVID-19 were identified in the infectious disease ward in Imam Khomeini hospital complex. The study also has been approved in ethics committee of Tehran University of medical sciences with ethic code: 99/11/101/16529. A blood sample was taken at the time of hospitalization and second one was taken 48 hours later. Blood samples are frozen at -80 ° C. After complete collection of samples, the ACE2 level of the samples was measured using serum sACE2 detection ELISA kit.

## Data Analysis

Data were analyzed using SPSS 25 (IBM Corp). Descriptive statistical analysis was used to describe items included in the survey. Data were expressed as medians (interquartile ranges, IQRs) for continuous variables. For bivariate analysis, the Mann–Whitney U test or t-test was used for continuous variables, and the χ^2^ or Fisher’s exact test for categorical variables. Survival curves were developed using the Kaplan–Meier method with log-rank test. p < 0.05 was considered statistically significant when a two-tailed test was performed.

## Result

Among the 38 patients, the mean age was 64.13 years, 52.6% were male, 42% were PCR positive test and 39.5% was expired. The most common presenting symptoms were cough (80%), fever (75.5%), dyspnea (60.5%), myalgias (35.8%), diarrhea (20%), and nausea and vomiting (15%). To measure the strength of the association between patient’s symptoms and outcome, Phi and Cramer’s V test was performed. There is not any significant association or correlation between COVID-19 patient’s symptoms and outcomes.

Laboratory findings on hospital admission are shown in Table 2. Of all patients, there were not any significant differences between expired and discharged group. ACE serum level in common with White blood cells, C-reactive protein, Erythrocyte Sedimentation Rate, Urea and Creatinine were higher in expired group (not significantly, P-value > 0.05). In contrast, Platelets, Hemoglobin, liver function tests (ALT, AST, Alp), and direct bilirubin were higher in discharged group (not significantly, P-value > 0.05).

**Table – 1.** Demographic characteristics of patients

| Table – 1 Demographic characteristics of patients |  |  |  |
| --- | --- | --- | --- |
|  | N = 38 |  |  |
| | Minimum | Maximum | Mean ± Std |
| **Age** | 30.00 | 88.00 | 64.13 ± 16.49 |
| **Mean Serum ACE Level** | 0.32 | 11.84 | 2.55 ± 2.41 |
| **Pulse Rate** | 58.00 | 120.00 | 86.39 ± 14.33 |
| **Respiratory Rate** | 18.00 | 40.00 | 24.89 ± 5.94 |
| **Systolic Blood Pressure** | 75.00 | 160.00 | 119.81 ± 19.42 |
| **Diastolic Blood Pressure** | 44.00 | 99.00 | 74.47 ± 11.66 |
| **White blood cells, 10<sup>3</sup>/L** | 3.50 | 27.80 | 10.61 ± 5.67 |
| **C–reactive protein, mg/dL** | 6.00 | 173.00 | 66.23 ± 42.08 |
| **Hemoglobin, g/dL** | 5.90 | 16.60 | 11.47 ± 2.61 |
| **Platelets, 10<sup>3</sup>/L** | 41.00 | 599.00 | 250.15 ± 137.30 |
| **Urea, mg/dl** | 23.00 | 230.00 | 80.00 ± 42.62 |
| **Creatinine, mg/dL** | 0.50 | 5.90 | 1.63 ± 1.32 |
| **ESR** | 10.00 | 139.00 | 61.07 ± 33.61 |
| **Blood Sugar, mg/dl** | 53.00 | 370.00 | 160.60 ± 79.50 |
| **Alanine aminotransferase, U/L** | 10.00 | 748.00 | 67.36 ± 119.04 |
| **Aspartate aminotransferase, U/L** | 13.00 | 270.00 | 57.52 ± 52.96 |
| **Alkaline Phosphatase** | 69.00 | 1939.00 | 295.73 ± 303.67 |
| **Total bilirubin, mg/dL** | 0.20 | 3.20 | 0.83 ± 0.54 |
| **Direct bilirubin, mg/dL** | 0.20 | 6.70 | 0.60 ± 1.03 |
| **O2 Saturation** | 75.00 | 95.00 | 87.81 ± 6.00 |
| **PH** | 7.24 | 7.63 | 7.39 ± 0.09 |
| **PCO2** | 16.10 | 67.70 | 40.77 ± 11.61 |
| **PT** | 12.00 | 47.30 | 16.67 ± 6.01 |
| **PTT** | 25.00 | 55.00 | 33.78 ± 6.72 |
| **INR** | 0.18 | 3.61 | 1.32 ± 0.49 |
| **Weight** | 49.00 | 116.00 | 74.42 ± 15.70 |
| **Hight** | 158.00 | 1162.00 | 193.13 ± 161.47 |

**Table – 2.**
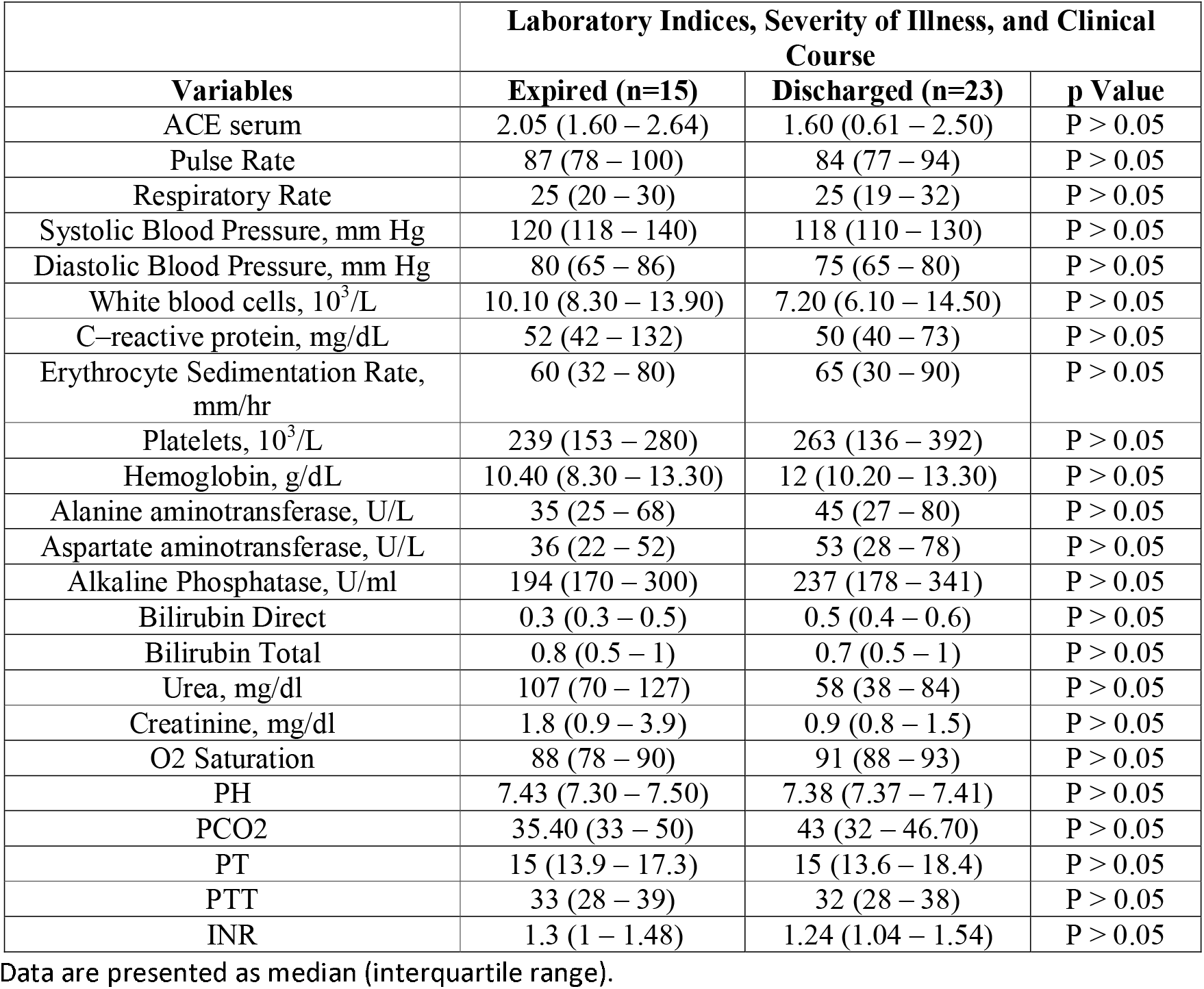
Laboratory Indices, Severity of Illness, and Clinical Course.

| **Table – 2** | **Laboratory Indices, Severity of Illness, and Clinical Course** |  |  |
| --- | --- | --- | --- |
| **Variables** | **Expired (n=15)** | **Discharged (n=23)** | **p Value** |
| ACE serum | 2.05 (1.60 – 2.64) | 1.60 (0.61 – 2.50) | P > 0.05 |
| Pulse Rate | 87 (78 – 100) | 84 (77 – 94) | P > 0.05 |
| Respiratory Rate | 25 (20 – 30) | 25 (19 – 32) | P > 0.05 |
| Systolic Blood Pressure, mm Hg | 120 (118 – 140) | 118 (110 – 130) | P > 0.05 |
| Diastolic Blood Pressure, mm Hg | 80 (65 – 86) | 75 (65 – 80) | P > 0.05 |
| White blood cells, 10<sup>3</sup>/L | 10.10 (8.30 – 13.90) | 7.20 (6.10 – 14.50) | P > 0.05 |
| C-reactive protein, mg/dL | 52 (42 – 132) | 50 (40 – 73) | P > 0.05 |
| Erythrocyte Sedimentation Rate, mm/hr | 60 (32 – 80) | 65 (30 – 90) | P > 0.05 |
| Platelets, 10<sup>3</sup>/L | 239 (153 – 280) | 263 (136 – 392) | P > 0.05 |
| Hemoglobin, g/dL | 10.40 (8.30 – 13.30) | 12 (10.20 – 13.30) | P > 0.05 |
| Alanine aminotransferase, U/L | 35 (25 – 68) | 45 (27 – 80) | P > 0.05 |
| Aspartate aminotransferase, U/L | 36 (22 – 52) | 53 (28 – 78) | P > 0.05 |
| Alkaline Phosphatase, U/ml | 194 (170 – 300) | 237 (178 – 341) | P > 0.05 |
| Bilirubin Direct | 0.3 (0.3 – 0.5) | 0.5 (0.4 – 0.6) | P > 0.05 |
| Bilirubin Total | 0.8 (0.5 – 1) | 0.7 (0.5 – 1) | P > 0.05 |
| Urea, mg/dl | 107 (70 – 127) | 58 (38 – 84) | P > 0.05 |
| Creatinine, mg/dl | 1.8 (0.9 – 3.9) | 0.9 (0.8 – 1.5) | P > 0.05 |
| O2 Saturation | 88 (78 – 90) | 91 (88 – 93) | P > 0.05 |
| PH | 7.43 (7.30 – 7.50) | 7.38 (7.37 – 7.41) | P > 0.05 |
| PCO2 | 35.40 (33 – 50) | 43 (32 – 46.70) | P > 0.05 |
| PT | 15 (13.9 – 17.3) | 15 (13.6 – 18.4) | P > 0.05 |
| PTT | 33 (28 – 39) | 32 (28 – 38) | P > 0.05 |
| INR | 1.3 (1 – 1.48) | 1.24 (1.04 – 1.54) | P > 0.05 |

Data are presented as median (interquartile range).

## Discussion

Our study was conducted to examine the relationship between serum ACE 2 level and COVID-19 patients’ prognosis. Coronavirus cell receptor is ACE2 [22] and several supposed treatments affect ACE2 receptor such as Chloroquine Hydroxychloroquine which block ACE2 receptors therefore limit virus availability to ACE2 receptors. It was also proposed that angiotensin-converting enzyme inhibitors (ACEIs) and angiotensin II receptor blockers (ARBs) might increase levels of ACE2. However, recent study reported these medications does not raise ACE2 levels in serum [23]. Accordingly, we proposed that, soluble ACE2 levels might correlated with severity of disease and patient’s prognosis. We found not any association between serum ACE2 level and patient outcome and severity. It could be because there was no correlation or due to limitation of our sample size.

In the present study, we did not find a significant difference in the serum ACE2 levels of the two groups of patients, which is aligned with similar studies [17,18]. However, we find that serum ACE 2 level was rise in patients with severe disease, who finally expired. Some studies hypothesized that, A significant increase of serum ACE2 activity may act as an endogenous nonspecific protective mechanism against SARS-CoV-2 infection that preceded the recovery of patients [19]. A study by Emilsson et al [20], suggested that upregulation of ACE2 levels may reflects severity of outcome in COVID-19. Besides, it seems that serum ACE activity on admission did not reflect disease severity [18]. Further studies are required for confirmation of association between serum ACE 2 and COVID-19 severity.

It was suggested that, Measurement of serum ACE2 antibody titers would help to identify who is likely to progress into acute respiratory distress, particularly in patients with cardiovascular disease and hypertension [21], Therefore, it might be possible to use serum ACE2 as indicator of ARDS.

## Conclusion

In summary, we did not find evidence for increased serum ACE2 (soluble) in discharged or expired patients with COVID-19. According to the results, no significant relationship was found between serum ACE2 level and patients’ prognosis, however, it seems that more research is needed. Due to study limitations, we suggest that serum ACE2 level determine in big sample size particularly in subgroup patients with diabetes, hypertension and asthma.

## Data Availability

Tehran University medical sciences.

## Acknowledgments

We would like to thank all medical staff for their effort in COVID-19 patient care.

